# A hierarchical clinical fusion transformer model for personalized opioid treatment: Development and validation in diabetic surgical patients

**DOI:** 10.64898/2026.06.04.26353331

**Authors:** Behzad Naderalvojoud, Brian J. Sutjiadi, Arman Koul, Catherine Curtin, Olivier Gevaert, Tina Hernandez-Boussard

**Affiliations:** Department of Medicine, Stanford University, Stanford, CA 94305, United States; Department of Surgery, Veterans Affairs Palo Alto Health Care System, Palo Alto, CA 94304, United States; Department of Biomedical Data Science, Stanford University, Stanford, CA 94305, United States

**Keywords:** Personalized opioid prescribing, HCF-Transformer, Machine learning, Postoperative opioid use, Diabetic surgical patients

## Abstract

**Background:** Machine learning (ML) models are increasingly used to predict adverse outcomes after surgery. However, most rely on static patient characteristics (e.g., age, comorbidities) and overlook clinician-controlled treatment decisions that can be actively modified at the point of care. Discharge opioid prescribing is a key modifiable, clinician-controlled decision, yet optimizing prescribing choices across multiple adverse outcomes remains underexplored in predictive modeling. This study addresses that gap by introducing a novel ML framework that explicitly separates fixed patient risk factors from modifiable prescribing options to support personalized, risk-informed opioid prescribing decisions.

**Methods:** We developed the Hierarchical Clinical Fusion Transformer (HCF-Transformer), an ML model designed to estimate patient-specific risks across four postoperative outcomes: prolonged opioid use (POU), chronic pain (CP), 30-day readmission, and opioid-associated outcomes (OAO). The model constructs patient risk profiles from fixed, non-modifiable baseline factors, followed by a transformer layer. Clinician-controllable discharge opioid regimens are modeled as alternative intervention candidates and fused with the fixed risk representation through a clinical fusion mechanism, enabling assessment and ranking based on predicted risks. A Total Relative Risk (TRR) metric, calibrated to each outcome prediction threshold, guides the recommendation process. We evaluated the model in diabetic surgical patients, a common high-risk population.

**Results:** The study included 157,853 unique diabetic surgical patients, with outcome prevalences ranging from 47.2% (POU) to 1.8% (OAO). The HCF-Transformer achieved the highest AUROCs, 0.798 for POU, 0.712 for 30-day readmission, 0.808 for CP, and 0.922 for OAO, outperforming Random Forest, FT-Transformer, and ResNet-based models. Compared to these baselines, HCF-Transformer generated more stable and discriminative risk estimates and demonstrated significant variation in TRR scores across discharge opioid options (ANOVA *p* < .01, η² > .01). This enabled consistent identification of lower-risk regimens tailored to patient-specific profiles.

**Conclusions:** The HCF-Transformer introduces a novel hierarchical fusion approach to optimize opioid prescribing by integrating static patient risk profiles with modifiable discharge options. Using transformer-based modeling and a quantifiable TRR metric, the model delivers personalized, risk-aware recommendations. This approach enables data-driven opioid prescribing tailored to individual risk and has the potential to improve postoperative outcomes in high-risk populations. Our findings demonstrate that integrating modifiable factors with structured risk profiles through a transformer-based fusion architecture can enhance decision-support systems, paving the way for more actionable and personalized AI in healthcare.

## 1. Introduction

Adverse postoperative outcomes remain a major challenge in surgical care, particularly those related to opioid prescribing [1,2]. Although machine learning (ML) models achieve strong performance in postoperative risk prediction, most rely on static patient characteristics, like comorbidities and demographics, and encode treatment decisions as fixed features [3,4]. As a result, they estimate risk only under the treatment actually rendered and cannot evaluate how alternative clinician-controlled decisions may influence predicted outcomes. Their architectures lack the capacity to incorporate modifiable clinical decisions—particularly opioid prescribing choices—as actionable inputs. This limitation constrains their utility for personalized decision support, where clinicians must balance fixed patient risk factors with modifiable treatment choices.

In this study, we define modifiable factors as clinician-controlled treatment decisions that can be actively adjusted for an individual patient. Discharge opioid prescribing represents a concrete and actionable decision point. We therefore architecturally separate a structured patient risk profile—derived from fixed clinical characteristics—from prescribing options, enabling systematic comparison of predicted risks across alternative regimens for the same patient. Rather than estimating causal effects, this work frames postoperative medication prescribing as a predictive decision-support problem: given a patient’s preoperative risk profile, how do different medication prescribing strategies compare in their associated risks? This perspective shifts predictive models from passive risk calculators toward actionable, risk-informed clinical tools.

Recent work demonstrates that deep learning (DL) architectures can achieve state-of-the-art performance in predicting postoperative complications when trained on static patient data [5,6]. These models typically learn joint representations of baseline patient characteristics and treatment variables to optimize overall risk prediction. While effective for capturing population-level associations, this approach embeds clinician-controlled treatment decisions within the same latent representation as fixed patient risk factors. Consequently, predicted risk reflects correlations observed in the training data rather than a structure designed to support systematic within-patient evaluation of alternative treatment strategies [7,8]. Without an explicit mechanism to separate modifiable decisions from underlying patient risk profile, these models remain oriented toward passive risk estimation rather than enabling actionable, risk-informed clinical decision support.

Addressing these limitations requires model architectures that represent patient risk profile and clinician-controlled treatment decisions as distinct yet interacting components within prediction. Fusion-based learning provides a principled strategy for integrating heterogeneous information while preserving its structural role. In clinical deep learning, fusion has been primarily used to combine multiple data modalities—such as medical imaging and structured clinical data—to improve diagnostic and prognostic performance [9–11], typically by merging modality-specific representations into a unified latent space [12–14]. However, existing fusion frameworks are not designed to support structured comparison of alternative treatment decisions within the same patient.

To address this gap, we developed the Hierarchical Clinical Fusion (HCF) Transformer, a novel architecture that performs fusion at the decision level within a single EHR-derived modality. Specifically, the model uses a transformer to learn a patient-specific risk profile from EHR-based clinical features—including demographics, comorbidities, prior opioid exposure, and physiological indicators—which is then fused with clinician-controllable discharge opioid prescribing options. By integrating modifiable prescribing choices with fixed patient risk representations, this approach enables within-patient comparison of alternative discharge opioid regimens, a capability that traditional ML models using flattened feature representations typically lack, and supports personalized, risk-informed opioid prescribing in high-risk surgical populations.

## 2. Methods

### 2.1. Data

This study analyzed diabetic patients aged 18–89 within the Veterans Health Administration (VHA), the largest integrated healthcare system in the United States, using national administrative and clinical data from 2008 to 2022. We included patients who underwent major surgeries and received an opioid prescription within 30 days before or after surgery. Surgeries were identified by the International Classification of Diseases (ICD) 9 and 10 and Current Procedural Terminology (CPT) codes, consistent with prior work [1]. The surgeries were categorized into 19 major surgery types (Supplementary Table S1) using the Clinical Classifications Software for Services and Procedures [15]. The opioid prescriptions were identified using 9 ingredient opioids listed in Supplementary Table S2. Patients were included if they had alanine aminotransferase (ALT), aspartate aminotransferase (AST), and glycated hemoglobin (A1C) measurements within six months preoperatively, at least two clinical visits before and after surgery, and survived for at least one year postoperatively. The one-year survival requirement was applied to ensure adequate follow-up for outcome ascertainment and complete capture of postoperative events. To isolate the impact of postoperative outcomes, patients who underwent any secondary surgery between 61 and 365 days after their initial surgery were excluded.

The study focused on four primary postoperative outcomes, each modeled as a binary prediction task. Prolonged opioid use (POU) was defined as any new opioid prescription between 90 and 365 days after discharge, based on a curated list of 9 RxNorm opioid ingredients used in prior work [15]. Opioid-related adverse outcomes (OAO) were defined as the occurrence of any diagnosis related to opioid overdose, abuse, or misuse between 1 and 365 days post-discharge, identified using ICD codes from prior work [2] and mapped to equivalent OMOP concept identifiers (Supplementary Table S3). Chronic pain (CP) was defined as the presence of a chronic pain diagnosis between 90 and 365 days after discharge, using the SNOMED diagnosis code (82423001) and its descendants. The 30-day readmission was defined as any emergency department visit or inpatient admission occurring within 30 days of discharge. For all outcomes, patients were labeled as positive (y = 1) if the corresponding event occurred within the defined time window and negative (y = 0) otherwise.

### 2.2. HCF-Transformer model

The overview of the prediction approach and architecture of the HCF-Transformer model, along with the approach for ranking discharge opioid options, are illustrated in Figures 1 and 2. Model inputs were organized into baseline and decision-related features (Figure 1). Baseline features consisted of a structured risk profile represented as five feature groups (11 features each; 55 total) capturing physiological, comorbidity, and medication-related factors available prior to prediction; these were treated as fixed, non-modifiable inputs to the transformer encoder (Supplementary Tables S4 and S5). Clinician-controlled treatment decision features—including surgical type (19 features) and discharge opioid category (6 features)—were incorporated during the hierarchical fusion stage. Together, these inputs comprised 80 features. Because these variables may vary at the time of prediction, this design enables patient-specific risk estimation under alternative clinician-controlled treatment scenarios. The details of the model architecture and the ranking approach are described in the following sections.

**Figure 1.**
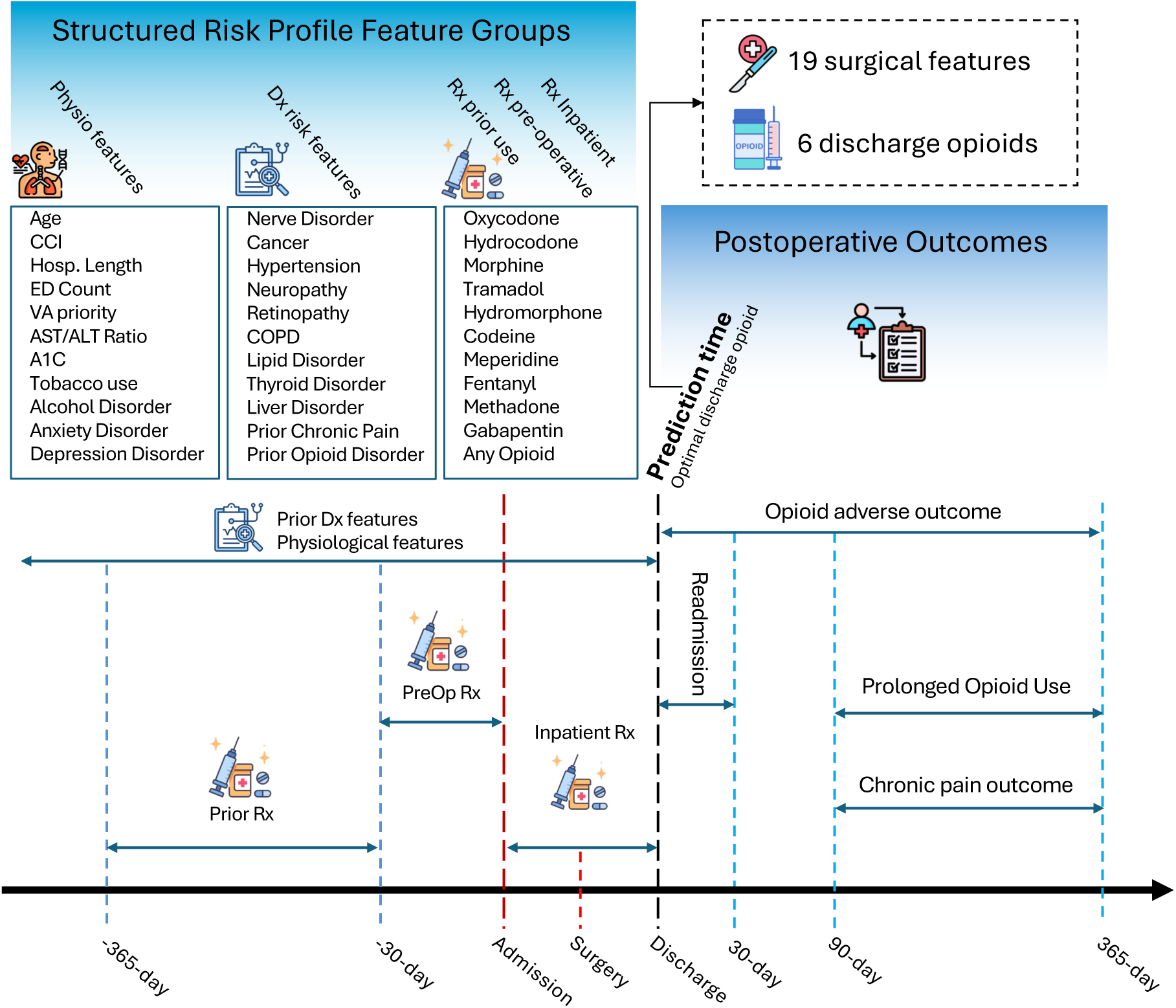
Overview of the prediction framework with structured risk profile feature groups and postoperative outcomes.

**Figure 2.**
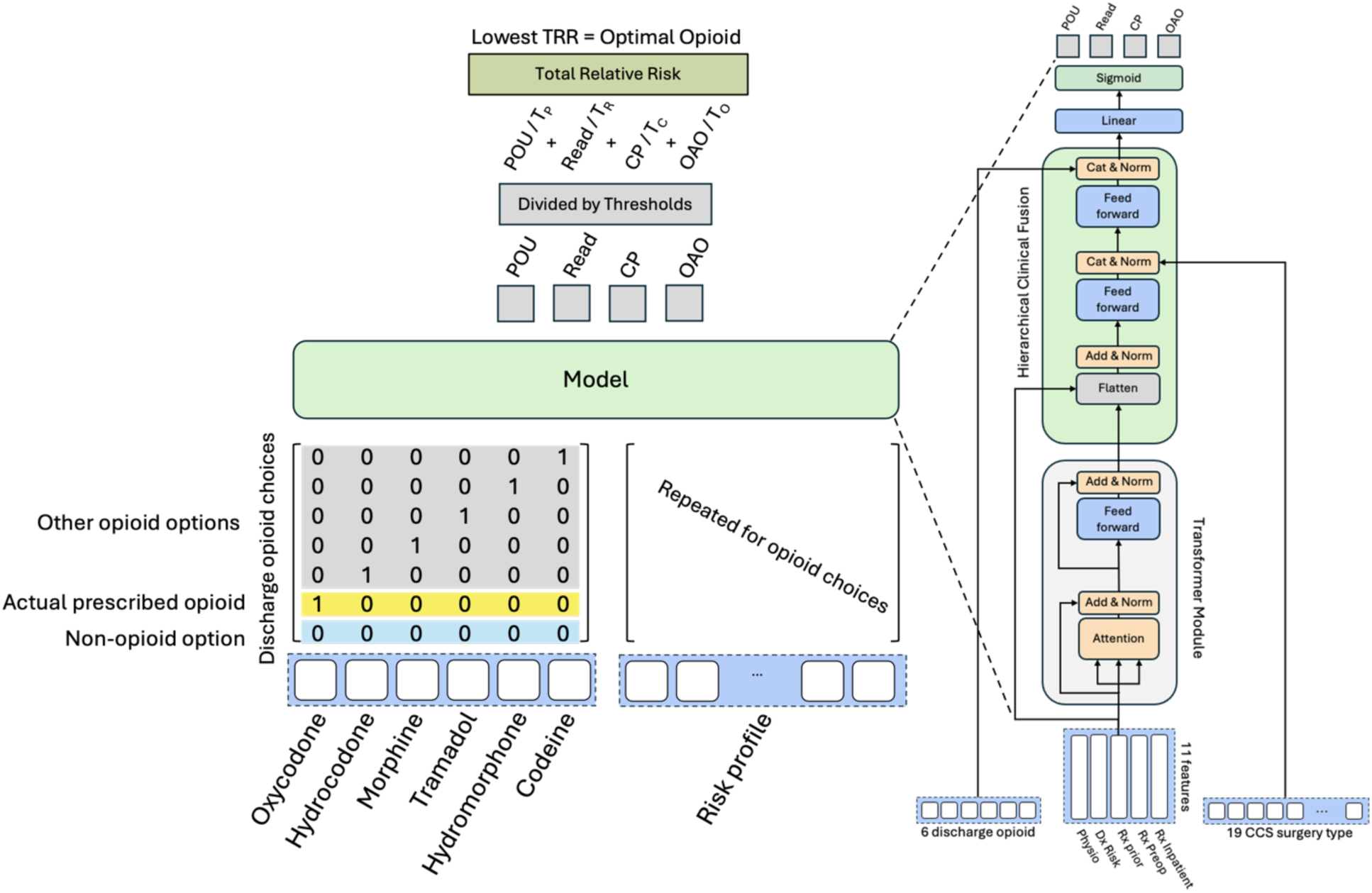
Overall architecture of HCF-Transformer and opioid ranking approach. POU: Prolonged opioid use; Read: 30-day readmission; CP: Chronic pain; OAO: Opioid-related adverse outcome; TP: Threshold of POU; TR: Threshold of Read; TC: Threshold of CP; TO: Threshold of OAO; TRR: Total relative risk.

Overview of the prediction framework, including input features and postoperative outcomes. The prior risk profile is organized into five feature groups (11 features each; 55 total): (1) physiological features (e.g., age, laboratory values, mental health, substance use history), (2) comorbidity/diagnosis-based features, and (3–5) medication-based features defined across three exposure windows (365 to 31 days before hospitalization, 30 to 1 days before hospitalization, and during hospitalization). In addition, 19 surgical-type features and 6 discharge opioid-type features are included, resulting in 80 total input features in the flattened representation. Four postoperative outcomes were predicted. See Supplementary Tables S4 and S5 for detailed code and temporal definitions.

#### 2.2.1. Transformer layer with patient-specific risk profile

We developed the HCF-Transformer model using a risk profile input with 11 features across five steps followed by transformer layer. The first step includes physiological features such as age, A1C, and the AST/ALT ratio. The second step captures prior diagnostic risk factors, such as hypertension, neuropathy, and liver disorders. The last three steps represent opioid and gabapentin use across three timeframes: prior use (365-31 days before surgery), preoperative use (30-1 days before surgery), and inpatient use (from admission to discharge). The details of diagnostic periods have been summarized in Figure 1. A clinical fusion layer incorporated surgical procedure types and discharge opioid options into the transformer output. We only considered six opioids when identifying discharge prescriptions, which are commonly used in outpatient settings: oxycodone, hydrocodone, morphine, tramadol, hydromorphone, and codeine [16,17].

#### 2.2.2. Hierarchical clinical fusion

The hierarchical clinical fusion (HCF) layer integrates contextual clinical features with modifiable treatment decisions in a structured manner. Rather than flattening all inputs into a single feature space, the hierarchy enables sequential conditioning in which upstream clinical context (e.g., surgical procedure type) informs downstream clinician-controllable decisions such as discharge opioid selection. This design reflects clinical decision-making workflows and supports scalable incorporation of additional modifiable components.

The fusion layer operates through three sequential modules, as shown in Figure 2. First, the transformer output is combined with its original input via a residual connection to preserve upstream representations and stabilize training, similar to residual networks (ResNets) [18]. The resulting tensors are flattened, summed, and normalized before entering the next fusion module. In the second module, the transformer representation is passed through a feedforward layer with an exponential linear unit (ELU) activation [19], producing a 20-neuron vector. This vector is concatenated with the 19 surgical features and normalized, allowing procedure-level context to condition the intermediate representation.

In the third module, the resulting 49-neuron vector is processed through another feedforward layer that generates a 12-neuron output. This representation is then concatenated with the 6 discharge opioid features and normalized, enabling treatment-specific information to refine the final representation. In the third module, the resulting 49-neuron vector is processed through another feedforward layer that generates a 12-neuron output. This representation is then concatenated with the 6 discharge opioid features and normalized, enabling treatment-specific information to refine the final representation. Finally, the fused vector is subsequently passed to a linear layer with four equally weighted sigmoid activations to generate four outcome probabilities. This hierarchical structure supports extensible decision-level fusion while reducing the risk that modifiable treatment features are diluted within a flattened representation.

#### 2.2.3. Total relative risk and optimal opioid prescribing

To support counterfactual evaluation of discharge opioid options, we estimate outcome risks under multiple prescribing scenarios, including the actual prescribed opioid, a non-opioid option, and alternative opioid classes (Figure 2). For each option, the model produces predicted probabilities for four outcomes. We define the Total Relative Risk (TRR) as the sum of normalized risks across outcomes, where each predicted risk is divided by a pre-specified outcome-specific threshold. Formally, for option 𝑘, given four outcomes:

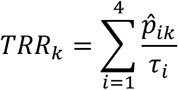

where *p̂_ik_* denotes the predicted probability of outcome 𝑖 under option 𝑘, and 𝜏_*i*_is the corresponding threshold. Outcome-specific thresholds serve as reference boundaries that contextualize predicted risks relative to clinically or operationally meaningful points, rather than relying on absolute probability values. By normalizing each predicted risk by its corresponding threshold, the resulting ratio indicates whether the estimated risk is below, near, or above a predefined decision boundary for that outcome. This normalization facilitates interpretation at the individual outcome level and mitigates the influence of differences in baseline outcome prevalence when aggregating risks. While these normalized risks are interpretable individually, their summation into TRR is intended as a composite decision score for comparing alternative prescribing options within the same patient, rather than as a directly interpretable probabilistic quantity. This design prioritizes relative, within-patient decision-making across actionable options over absolute risk estimation. In this study, thresholds were selected based on validation performance criteria, such as maximizing F1 score, but can be adjusted to reflect institutional priorities or alternative risk tolerances. The discharge option with the lowest TRR is considered the most favorable relative to available alternatives under the model’s aggregated, normalized predicted risk scores across outcomes.

### 2.3. Model training and evaluation

The cohort was split into training, validation, and test sets using a stratified approach based on the outcome. Specifically, 80% of the cohort was used for training and validation, with 90% assigned for training and 10% for validation. The remaining 20% of the cohort served as the test set. The best model was selected based on the area under the receiver operating characteristic curve (AUROC), and outcome thresholds were determined according to the best F1 scores obtained from the validation set for each outcome. Hyperparameters were tuned using a grid search strategy, resulting in the following best parameters: the model’s transformer was configured with one head and one layer; the dimension of the feedforward layer was set to 512, and a dropout rate of 0.1 was used. The optimizer was AdamW [20], with a weight decay of 1e-6. The learning rate was initialized at 0.001 and decreased by a factor of 0.1 after a plateau was observed for 5 epochs.

We compared our approach with Random Forest (RF) as well as ResNet [18] and Feature Tokenizer + Transformer (FT-Transformer) [21], both widely used in tabular modeling studies [22–24]. The same grid search strategy was employed to identify the best hyperparameters for each model. For the Random Forest, the parameters were configured as follows: the number of estimators was set to 600, the maximum depth was limited to 50, the minimum samples required to split was set to 3, and the minimum samples required in a leaf node was set to 4. For ResNet and FT-Transformer, we used an embedding dimension of 32, along with hyperparameters like those utilized in the HCF-Transformer.

The performances of the models were evaluated using AUROC and the area under precision-recall curve (AUPRC). Additionally, F1 scores were calculated at specific thresholds to facilitate comparisons between the models. We conducted a one-way ANOVA to assess whether TRRs predicted by models are significantly different across the opioid options. To quantify the magnitude of the observed differences, we calculated the effect size using η² (eta-squared) [25]. In addition, we estimated 95% confidence intervals for the TRR values using the standard error and the corresponding z-score.

## 3. Results

Among 157,853 surgical diabetic patients, 47.2% had POU, 9.2% had 30-day readmission, 10.4% had CP, and 1.8% had OAO (Table 1). Most were male (97.5%), white (81.7%), and non-Hispanic (95.2%), with an average age of 65.1 (SD=9.16). We trained the models on 113,653 patients, validated them on 12,629 patients, and tested them on 31,571. All the following results reflect the model outputs on the test set.

**Table 1.** Cohort characteristics with respect to study outcomes.

| Characteristics | All (N=157,853) |  |  |  |
| --- | --- | --- | --- | --- |
|  | POU<br>(N=74,466)<br>47.2% | 30-Day-Read.<br>(N=14,458)<br>9.2% | CP<br>(N=16,369)<br>10.4% | OAO<br>(N=2,778)<br>1.8% |
| **Age** |  |  |  |  |
| Median (1st Qu, 3rd Qu)<br>[Mean] | 64.00 (59.00,<br>70.00) [64.20] | 66.00 (60.00,<br>72.00) [65.86] | 64.00 (58.00,<br>70.00) [63.71] | 61.00 (56.00,<br>66.00) [60.43] |
| **Gender** |  |  |  |  |
| Female | 3910 (5.3%) | 609 (4.2%) | 1306 (8.0%) | 166 (6.0%) |
| Male | 70556 (94.7%) | 13849 (95.8%) | 15063 (92.0%) | 2612 (94.0%) |
| **Race** |  |  |  |  |
| White | 59155 (79.4%) | 11584 (80.1%) | 12907 (78.9%) | 1951 (70.2%) |
| Asian | 270 (0.4%) | 70 (0.5%) | 61 (0.4%) | 5 (0.2%) |
| Black | 13722 (18.4%) | 2589 (17.9%) | 3140 (19.2%) | 775 (27.9%) |
| AI/AN | 733 (1.0%) | 115 (0.8%) | 141 (0.9%) | 31 (1.1%) |
| Hawaiian | 586 (0.8%) | 100 (0.7%) | 120 (0.7%) | 16 (0.6%) |
| **Ethnicity** |  |  |  |  |
| Non-Hispanic | 69375 (93.2%) | 13410 (92.8%) | 15319 (93.6%) | 2601 (93.6%) |
| Hispanic | 5091 (6.8%) | 1048 (7.2%) | 1050 (6.4%) | 177 (6.4%) |
| **Any prior opioid use** |  |  |  |  |
| Opioid-Naive | 18406 (24.7%) | 5809 (40.2%) | 4400 (26.9%) | 585 (21.1%) |
| Opioid-Exposed | 56060 (75.3%) | 8649 (59.8%) | 11969 (73.1%) | 2193 (78.9%) |
| **Hospitalization Length** |  |  |  |  |
| Median (1st Qu, 3rd Qu)<br>[Mean] | 3.00 (0.00, 8.00)<br>[5.69] | 6.00 (2.00, 11.00)<br>[8.38] | 3.00 (0.00, 7.00)<br>[5.52] | 4.00 (0.00, 8.00)<br>[6.46] |
| **Prior ED visits** |  |  |  |  |
| Median (1st Qu, 3rd Qu)<br>[Mean] | 3.00 (0.00, 8.00)<br>[6.50] | 3.00 (1.00, 9.00)<br>[7.86] | 4.00 (1.00, 12.00)<br>[9.63] | 7.00 (2.00, 17.00)<br>[15.02] |
| **VA priority group norm** |  |  |  |  |
| Median (1st Qu, 3rd Qu) | 1.00 (0.20, 1.00) | 1.00 (0.20, 1.00) | 1.00 (0.25, 1.00) | 1.00 (0.20, 1.00) |
| [Mean] | [0.68] | [0.64] | [0.71] | [0.66] |
| **CCI** |  |  |  |  |
| Median (1st Qu, 3rd Qu) | 5.00 (3.00, 7.00) | 6.00 (4.00, 8.00) | 6.00 (4.00, 8.00) | 6.00 (4.00, 8.00) |
| [Mean] | [5.53] | [6.16] | [5.99] | [6.17] |
| **A1C** |  |  |  |  |
| Median (1st Qu, 3rd Qu) | 6.80 (6.10, 7.80) | 6.90 (6.20, 8.00) | 6.70 (6.09, 7.70) | 6.60 (5.90, 7.80) |
| [Mean] | [7.12] | [7.29] | [7.08] | [7.10] |
| **AST/ALT Ratio** |  |  |  |  |
| Median (1st Qu, 3rd Qu) | 0.93 (0.73, 1.18) | 0.96 (0.76, 1.23) | 0.92 (0.72, 1.18) | 1.00 (0.76, 1.27) |
| [Mean] | [1.01] | [1.06] | [1.00] | [1.07] |
| Study outcomes: prolonged opioid use (POU), development of chronic pain (CP), 30-day readmission, and opioid-related adverse outcomes (OAO). |  |  |  |  |
| VA priority group: A classification used by the Veterans Health Administration to determine eligibility and cost-sharing based on service-connected disability, income level, and other factors. Ordinal variable scaled as the inverse of the priority group number so that higher values correspond to higher enrollment priority. |  |  |  |  |

All models were extensively tuned to achieve their best predictive performance, as the goal of this evaluation was not to maximize performance differences, but to assess whether models with comparable predictive performance can support within-patient comparison of alternative discharge opioid options. Figure 3 presents a comprehensive comparison of predictive performance between the proposed HCF-Transformer and baseline models (Random Forest and FT-Transformer) across four clinical outcomes: POU, 30-day readmission, CP, and OAO outcomes. Model performance was evaluated using both Precision-Recall (PR) and Receiver Operating Characteristic (ROC) curves with corresponding AUC values with 95% confidence intervals estimated via bootstrap resampling. Across outcomes, predictive performance was broadly comparable across models. The HCF-Transformer demonstrated similar AUROC and AUPRC values, with modest improvements observed for certain outcomes such as POU (AUROC 0.798 [95% CI: 0.793–0.803], AUPRC 0.795 [95% CI: 0.788–0.801]). The HCF-Transformer also showed smoother and more stable PR and ROC curves across thresholds, suggesting consistent discrimination over a wide operating range, particularly for POU. This behavior is important for clinical deployment, where threshold selection may vary by institutional priorities. Consistent with AUROC and AUPRC findings, precision, recall, and F1 scores were broadly similar across models (Supplementary Table S6), further supporting comparable predictive performance despite differences in model architecture.

**Figure 3.**
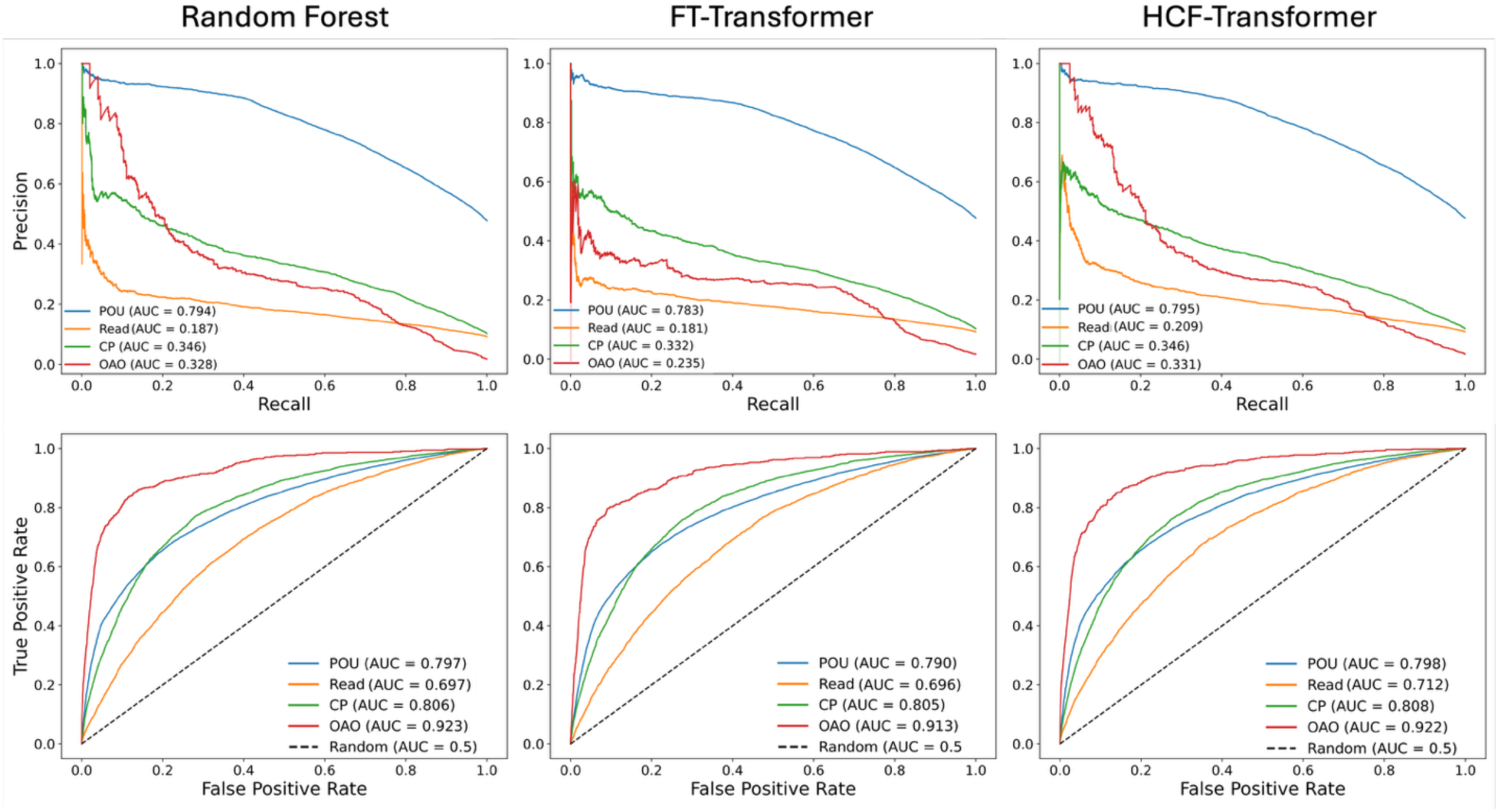
Model prediction performances.

Although predictive metrics were similar across models, discrimination alone does not fully characterize how models behave when used to compare clinician-controllable treatment options. We therefore examined model performance within a counterfactual clinical simulation to determine how architectural differences influence opioid risk stratification.

Model performances through Precision-Recall (PR) curves (top panel) and Receiver Operating Characteristic (ROC) curves (bottom panel) for the Random Forest, FT-Transformer, and HCF-Transformer models. POU: Prolonged opioid use; Read: 30-day readmission; CP: Chronic pain; OAO: Opioid-related adverse outcome

To this end, we trained the RF model by flattening the 80 features used in HCF-Transformer. Both models estimated the risk of outcomes and calculated TRR values for seven discharge opioid options, aiming to identify the lowest-risk choice for each patient. The TRR were calculated based on the thresholds achieving the highest F1 scores in the validation set. Figure 4 visualizes the results using a circular bar graph over the test set. The inner layer, closest to the center, represents the actual opioids prescribed, with percentages reflecting their prescription rates. The outer layer displays bar plots for the mean TRR values of the seven options, with the actual prescribed opioid highlighted.

**Figure 4.**
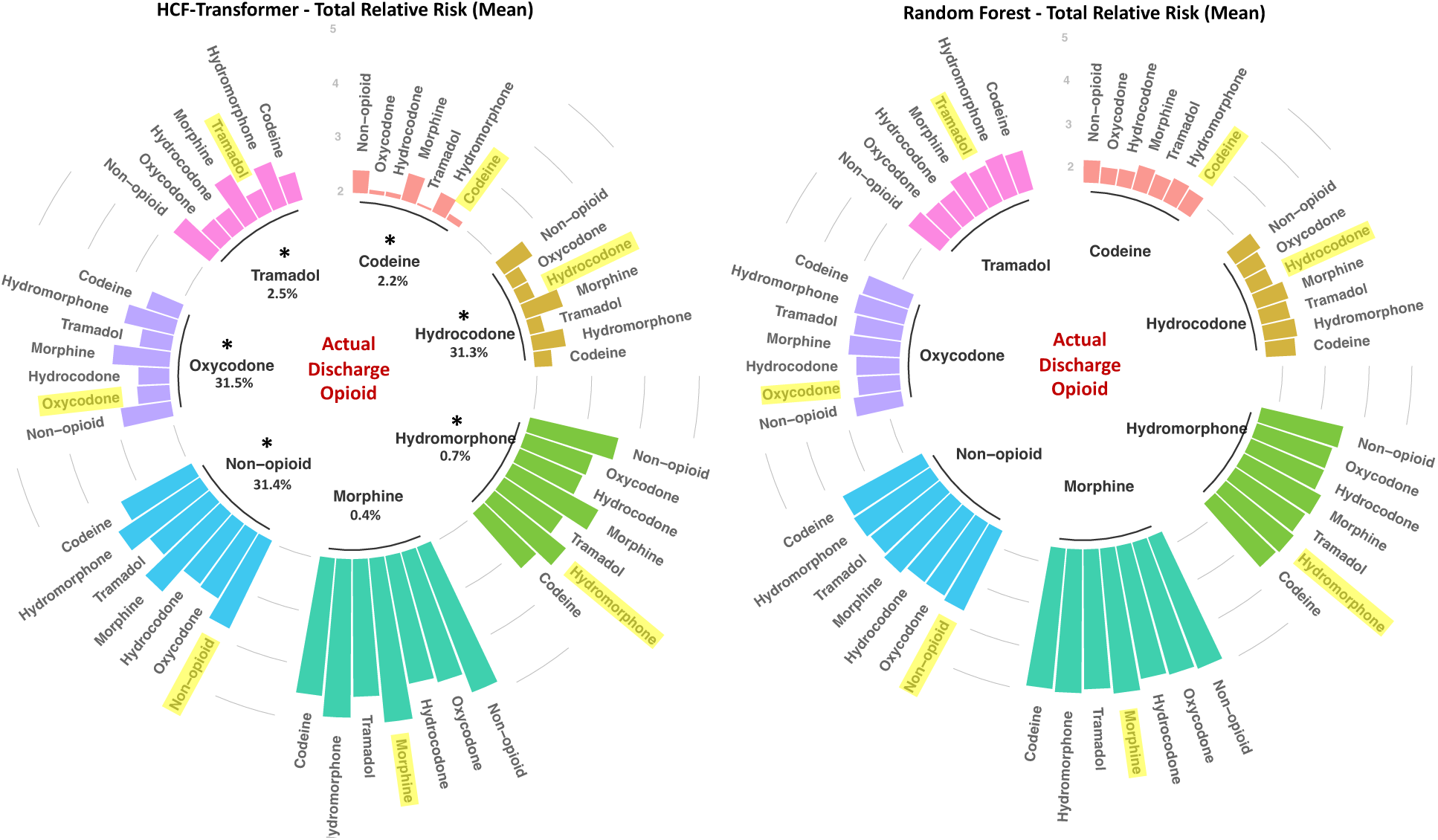
Total relative risks (TRR) of actual vs. alternative discharge opioids. Total relative risks (TRR) of actual vs. alternative discharge opioids based on Random Forest and HCF-Transformer models. * indicates ANOVA p < .01 with effect size η² > .01. Highlighted opioids represent TRRs for the actual prescribed discharge opioid.

Although both models achieved comparable AUCs, the figure reveals important differences in how they assess risk. The RF model produced relatively flat, undifferentiated TRR distributions, failing to capture clear differences between opioid alternatives. In contrast, the HCF-Transformer showed measurable differences across options (η² > .01), indicating non-trivial effect sizes, with statistical significance observed given the large sample (ANOVA p < .01). It consistently identified certain opioids as higher or lower risk, resulting in a clearer relative ranking.

We conducted a sensitivity analysis to evaluate the impact of varying outcome thresholds on TRR estimates derived from the HCF-Transformer model. Supplementary Figure S1 presents TRRs calculated using an alternative thresholding strategy aligned with outcome prevalences shown in Table 1. The overall TRR distributions were similar across thresholding approaches, with consistent identification of the three most favorable opioid options, as shown in Figure 4. This indicates that the relative comparisons were stable to reasonable variations in threshold selection.

We classified the opioid options into three categories: the actual opioids prescribed, non-opioids, and the optimal opioid choices as identified by both the RF and HCF-Transformer models. Figure 5 shows the distribution of TRRs for these three categories. The left violin plot for the RF model shows overlapping TRR distributions with minimal differentiation among actual, non-opioid, and optimal choices, indicating a limited ability to distinguish risk levels. In contrast, the right plot for the HCF-Transformer presents a much clearer separation, consistently highlighting that optimal opioids have lower TRRs compared to those actually prescribed. This pronounced differentiation highlights the HCF-Transformer’s ability to distinguish predicted relative risks across opioids, which may support more informed prescribing decisions.

**Figure 5.**
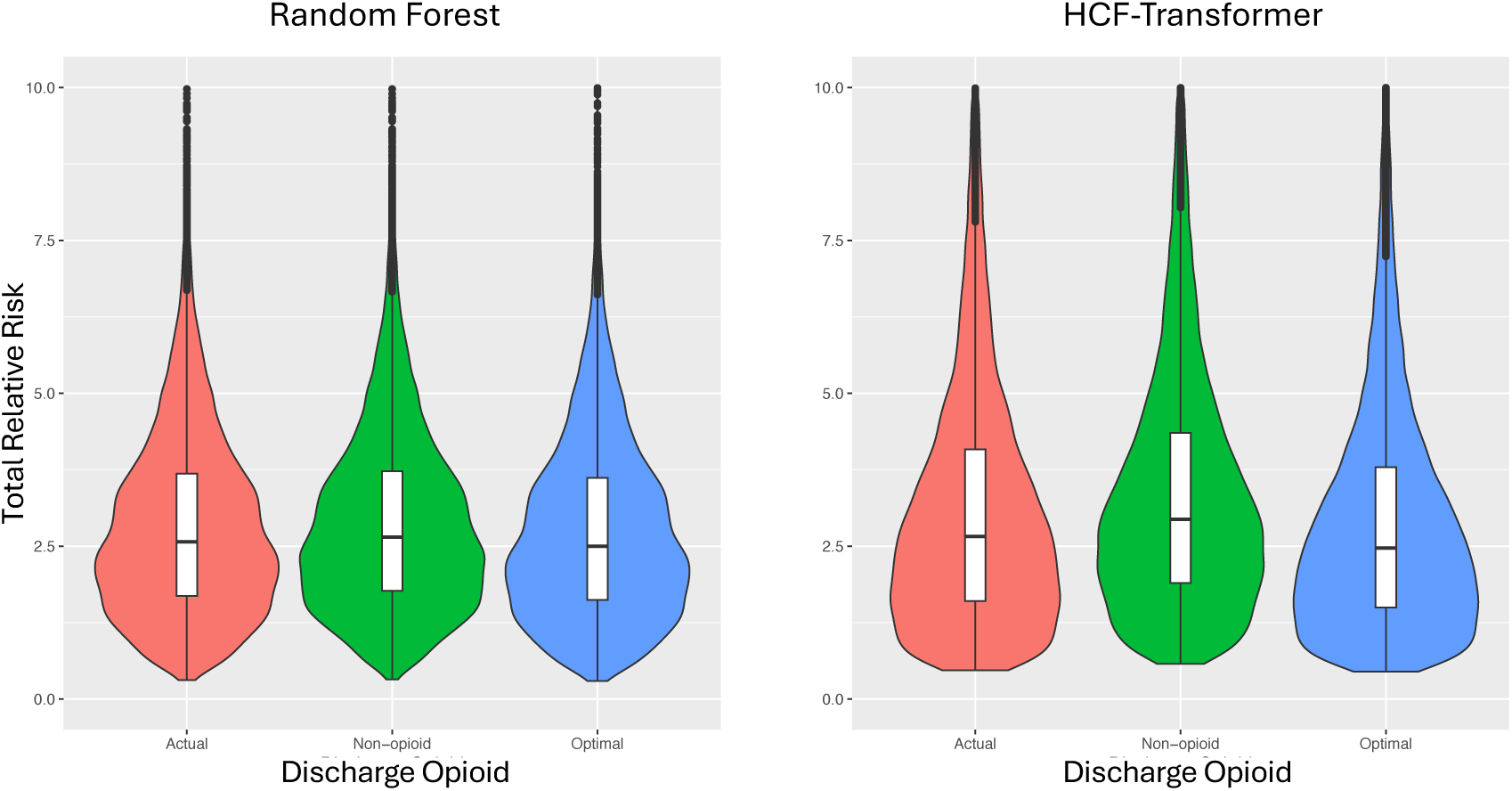
Distribution of total relative risk for actual opioid, non-opioid, and optimal opioid. Distribution of total relative risk for actual opioid prescriptions compared to non-opioid and optimal opioid options, as calculated by the Random Forest and HCF-Transformer models.

Figure 6 presents a comparison of patient characteristics associated with optimal discharge opioids as predicted by the RF and HCF-Transformer models. Each panel showcases different patient metrics, including age, A1C levels, AST/ALT ratio, and CCI, for various opioids identified as optimal. It is worth noting that oxycodone was optimal for only a few cases and excluded from the HCF-Transformer’s analysis as it was insufficient for statistical evaluation. This figure highlights important differences in how each model identifies optimal opioid options and the corresponding patient characteristics for each. Notably, the HCF-Transformer identified only three opioids as optimal, indicating a more selective approach, while the RF model identified six, suggesting a broader range of options as optimal. This overestimation in the RF model is reflected in the four patient characteristics groups, where the RF model shows unclear patterns across the various optimal opioids, making it difficult to distinguish between groups. Conversely, the HCF-Transformer leads to a clear distinction among patient groups for its selected optimal opioids. The details of differences between the average of TRRs for model-recommended opioids and actual opioids are given in the supplementary Table S7. This demonstrates HCF-Transformer’s strength in accurately characterizing the patient profiles’ association with specific opioid choices and its precision in guiding effective prescribing decisions based on patient characteristics.

**Figure 6.**
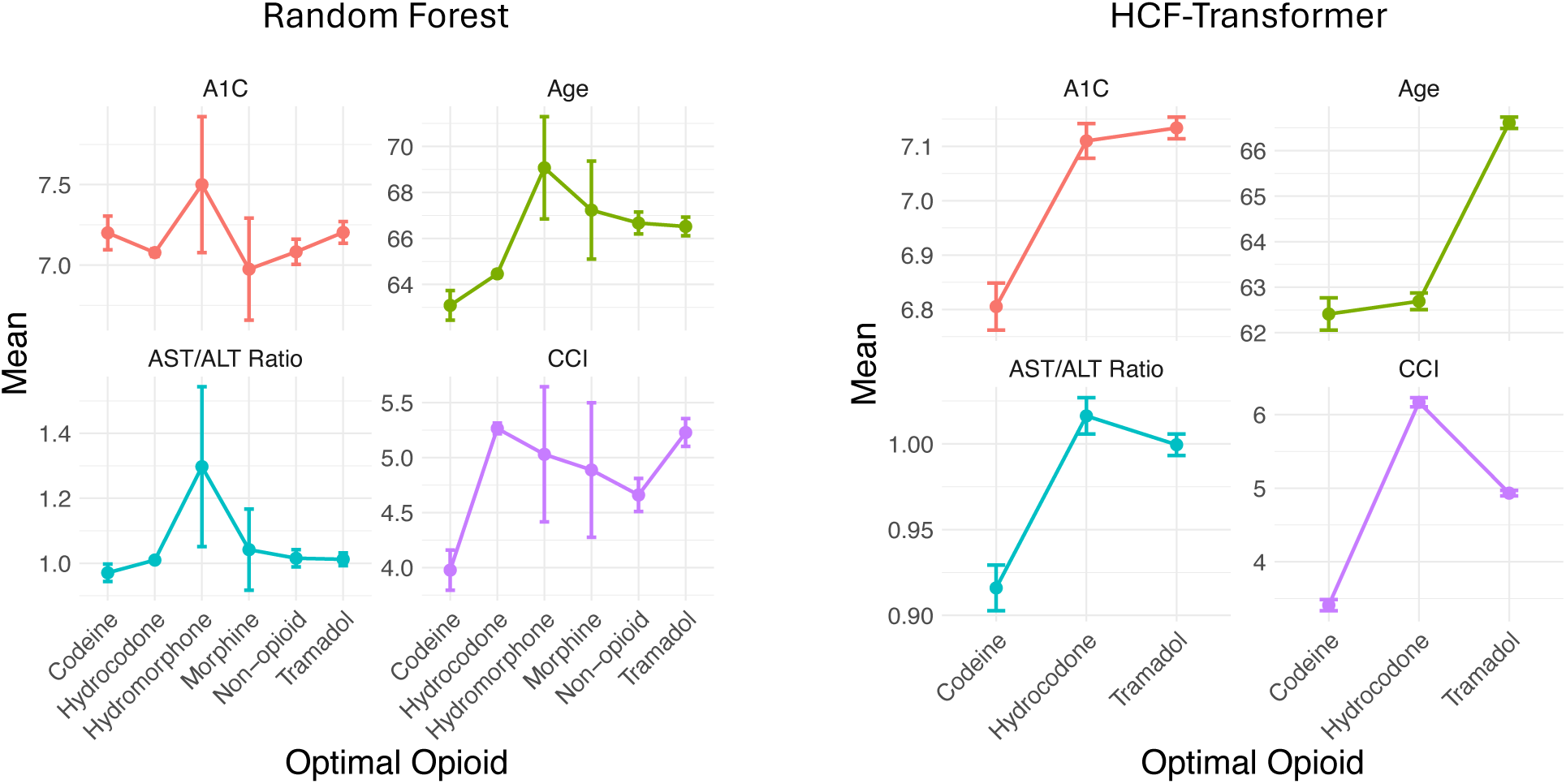
Characteristics of patients with optimal discharge opioids. Characteristics of patients with optimal discharge opioids predicted by the Random Forest and HCF-Transformer models. Each panel presents metrics, including A1C, age, AST/ALT ratio, and Charlson Comorbidity Index (CCI), corresponding to various optimal opioids.

## 4. Discussion

In this study, we developed and validated the HCF-Transformer model, an architecture designed to support within-patient evaluation of discharge opioid prescribing options. All comparator models were extensively tuned to achieve strong predictive performance, allowing the evaluation to focus not on marginal accuracy gains but on whether structurally different models can enable clinically meaningful comparison of alternative treatment options. Within this setting, the HCF-Transformer demonstrated that separating patient risk profiles from clinician-controlled prescribing decisions allows predicted risk to be examined across multiple regimens for the same patient. Rather than functioning solely as a risk prediction tool, this approach provides a structured mechanism to suggest medication regimens associated with the lowest risk of poor pain outcomes. By aligning predictive modeling with clinical decision points, the HCF-Transformer illustrates how decision-aware architectures can extend the role of machine learning from passive risk estimation toward more actionable forms of decision support in postoperative care.

### 4.1. Personalized risk estimation to inform opioid prescribing

The HCF-Transformer supports personalized risk estimation by generating patient-specific predicted risks across opioid options, facilitating comparison of total relative risk values. By structurally separating patient risk profiles from clinician-controlled prescribing decisions, the model supports evaluation of how predicted risk shifts under alternative regimens rather than anchoring estimates to the treatment historically observed. The TRR metric further enhances interpretability by translating outcome-specific predictions into a transparent summary of relative risk, allowing clinicians to quickly distinguish prescribing options associated with higher overall postoperative risk while still retaining visibility into individual complications. Instead of collapsing outcomes into a composite endpoint [2,26] or only identifying high-risk patients only after a prescription is made [27–30], our framework allows each outcome to be examined separately for every opioid option, making potential risk tradeoffs more transparent. Such structured comparison may better align with real-world prescribing, where clinicians must weigh multiple competing risks when selecting an analgesic strategy. Similar to interpretation of other ML model outputs, it is important to note that these rankings reflect model-based risk estimates rather than causal effects.

### 4.2. Predicted opioid-associated risk vs. actual prescribing

Although many ML models predict opioid risk outcomes, few evaluate how predicted risk varies across specific opioid options in a way that can inform prescribing decisions [3]. In this study, relative predicted risks across seven discharge opioid options were compared with observed prescribing patterns in diabetic surgical patients. Hydrocodone, tramadol, and codeine were associated with lower predicted risks, whereas oxycodone—the most frequently prescribed opioid—showed comparatively higher predicted risks, consistent with prior literature [31]. At the same time, tramadol has been linked to hypoglycemia in diabetic patients [32], underscoring that models focused on common postoperative outcomes, may not capture rare but clinically important adverse events. Similarly, the higher predicted risks observed for the non-opioid option, consistent with previous research [33], should be interpreted cautiously, as treatment selection in observational data may reflect confounding clinical factors. Accordingly, these results should not be interpreted as causal or prescriptive evidence of superiority of one opioid strategy over another. Rather, they illustrate how decision-aware models can reveal structured differences in predicted risk across prescribing options, thereby motivating further validation to assess their clinical reliability and generalizability.

Our findings also contribute to the broader discussion of what constitutes “optimal” opioid prescribing within ML frameworks, where optimality is typically defined through statistical prediction rather than causal clinical effects. Prior studies have approached this question from multiple angles, including reinforcement learning strategies to guide morphine dosing [34], optimization of prescription duration to reduce refills [35], and classification models predicting opioid receipt [36]. In contrast, our approach focuses on variation in predicted risk across alternative opioid agents—an underexplored dimension that emphasizes comparative risk rather than treatment assignment. Together, this highlights the multifaceted nature of prescribing decisions and suggests that decision-aware predictive models may complement existing strategies by making risk tradeoffs more explicit.

### 4.3. Optimal opioids vs. patient characteristics

We further estimated how model-estimated lower-risk opioid options varied across patient characteristics. Compared with the RF model, the HCF-Transformer demonstrated clearer associations between predicted lower-risk opioids and clinically relevant features such as age, glycemic control (A1C), comorbidity burden (CCI), and liver function indicators (AST/ALT ratio). This suggests that the hierarchical representation in HCF-Transformer may facilitate more structured characterization of how patient risk profiles interact with opioid options to shape predicted risk. Although only a subset of highly important features was examined, these findings indicate that decision-aware architectures can provide more clinically coherent stratification than models relying on flattened feature representations. This pattern was less apparent in RF, FT-Transformer, and ResNet models despite comparable predictive performance, underscoring that architectural design may influence the interpretability of within-patient comparisons even when overall accuracy is similar. Interpretability in this framework is defined at the decision level rather than through feature-level attribution. Unlike tree-based models, transformer-based hierarchical fusion does not yield directly comparable feature importance scores without post-hoc methods, and attention weights were not treated as explanations given prior evidence that attention is not a reliable proxy for importance [37–39]. Instead, interpretability arises from structured comparison of predicted risks across clinically actionable prescribing options using TRR-based evaluation. These findings highlight the potential importance of architectural design in tailoring opioid prescriptions for diabetic patients, where the risks associated with specific opioids [40,41] may vary based on individual characteristics.

### 4.4. Clinical implications and Limitations

The HCF-Transformer illustrates how decision-aware predictive modeling could be integrated into postoperative prescribing workflows. By enabling structured comparison of predicted relative risks across discharge opioid options, the model may support more transparent evaluation and more informed prescribing decisions. Importantly, these predictions are intended to complement, rather than replace, clinical judgment and should be interpreted as associations rather than causal effects. Beyond individual encounters, this framework highlights the potential value of incorporating patient-specific risk stratification into postoperative pain management strategies, particularly in high-risk populations such as diabetic surgical patients. More broadly, although this study focused on opioid prescribing, the hierarchical fusion framework is designed to structurally integrate patient risk profiles with clinician-controllable treatment decisions. This design may be adaptable to other clinical contexts where within-patient comparison of alternative interventions is needed, though further prospective validation is required.

For implementation, the model may be integrated as a decision-support tool at the time of discharge prescribing. In this study, outcome-specific thresholds (Supplementary Table S6) were defined using validation data to balance sensitivity and precision and were incorporated into the TRR framework. For deployment, similar validation-based thresholds may serve as a reasonable default, with adjustment according to institutional priorities. While thresholds affect absolute risk classification, they do not change the relative comparison of prescribing options within a patient, as supported by the sensitivity analysis across alternative threshold choices.

The limitations of this study should be considered when interpreting its findings. First, the data were derived from the VHA population, which is predominantly male and older, potentially limiting generalizability to more diverse healthcare settings. External validation in non-VHA systems will be necessary to evaluate performance stability and applicability across broader demographic and clinical contexts. Second, our model relies on administrative and structured EHR data, which may lack granularity for variables such as pain severity or patient-reported outcomes. While this is a common limitation in large-scale studies, future research could incorporate patient-reported data to enhance the model’s accuracy. Third, our analysis focused on opioid-related outcomes and did not model multimodal pain management strategies. While this reflects the current data availability, future work could explore the integration of multimodal pain management approaches. Fourth, model-identified lower-risk opioid options were defined based on four selected outcomes, which may not fully capture the range of clinically meaningful risks relevant to diabetic surgical patients. Future research could include more risk outcomes in the model to improve opioid prescribing. Fifth, formal evaluation of model calibration was not performed. Because TRR is derived from threshold-based outcome predictions within a unified framework, relative rankings may therefore be less sensitive to uniform probability shifts. However, assessment and potential recalibration of predicted probabilities would be necessary prior to clinical implementation to ensure accurate interpretation of absolute risk estimates. Finally, the requirement for one-year postoperative survival may introduce survivor bias by excluding patients with early mortality, potentially limiting applicability to individuals at highest short-term risk. Despite these limitations, our study provides a critical foundation for advancing personalized opioid prescribing in diabetic surgical patients.

## 5. Conclusion

The HCF-Transformer introduces a hierarchical fusion framework that supports within-patient comparison of discharge opioid options by structurally integrating patient risk profiles with clinician-controllable prescribing decisions. By combining transformer-based representation learning with TRR-based risk aggregation, the model enables interpretable comparison of predicted postoperative risks across alternative regimens while maintaining comparable predictive performance to established models. Rather than functioning solely as a passive risk calculator, this approach illustrates how predictive architectures can be designed to support structured decision-making at clinically relevant choice points. More broadly, our findings suggest that decision-aware model design may extend the role of clinical ML from outcome prediction toward more actionable forms of risk-informed decision support.

## Ethics approval and consent to participate

This study is a retrospective analysis of observational health data that has received approval from the Institutional Review Board (IRB) at Stanford University (Protocol #47644 and Protocol #34551). The research complied with the guidelines outlined in the Helsinki Declaration. The data was anonymized and pooled prior to access and analysis, so the informed consent of participants was waived by the Stanford University IRB.

## Availability of data and materials

The dataset used in this study is not publicly available due to ethical restrictions on patients’ personal health data. However, they can be regenerated using the source codes available at the GitHub repository https://github.com/su-boussard-lab/HCF-Transformer for any OMOP CDM database, including the VHA OMOP data used in this study. VHA OMOP access can be requested through DART for research projects. For more information on access visit https://www.herc.research.va.gov/include/page.asp?id=omop

## Authors’ contributions

THB takes responsibility for the integrity of the data and the accuracy of the data analysis. Concept and design: THB; Methodology: BN and THB; Statistical analysis: BN; Collection of data: BN; Investigation and validation: BN, BS, AK; Interpretation of data: All authors; Drafting of the manuscript: BN; Critical revision of the manuscript: All authors; Administrative, technical, or material support: THB; VA resources: CC; Study supervision: THB

## Data Availability

The dataset used in this study is not publicly available due to ethical restrictions on patient personal health data. However, they can be regenerated using the source codes available at the GitHub repository https://github.com/su-boussard-lab/HCF-Transformer for any OMOP CDM database, including the VHA OMOP data used in this study. VHA OMOP access can be requested through DART for research projects. For more information on access visit https://www.herc.research.va.gov/include/page.asp?id=omop

