## Supplementary Materials for "A hierarchical clinical fusion transformer model for personalized opioid treatment: Development and validation in diabetic surgical patients"

**Supplementary Table S1.** Surgical procedure types used in the study, along with their CCS procedure categories.

| **Surgery types** | **Multi-Level CCS Procedure Categories** |
|---|---|
| Appendectomy | 9.12 |
| Coronary artery bypass graft (CABG) | 7.2 |
| Colorectal resection | 9.10 |
| Distal radius fracture* | 14.3 |
| Excision; lysis peritoneal adhesions | 9.22 |
| Hysterectomy; abdominal and vaginal | 12.5 |
| Inguinal and femoral hernia repair | 9.17 |
| Knee replacement* | 14.7 |
| Oophorectomy; unilateral and bilateral | 12.1 |
| Therapeutic procedures on joints | 14.15 |
| Partial excision bone | 14.1 |
| Spinal fusion | 14.11 |
| Treatment; fracture or dislocation of hip and femur* | 14.3 |
| Treatment; fracture or dislocation of lower extremity* | 14.3 |
| Cholecystectomy and common duct exploration | 9.16 |
| Laminectomy; excision intervertebral disc | 1.3 |
| Mastectomy* | 15.1 |
| Open prostatectomy | 11.2 |
| Thoracotomy* | 6.8 |

* Type of surgery corresponding to a subset of CCS procedure category.

CCS (Clinical Classifications Software) category names are available at:
https://hcup-us.ahrq.gov/toolssoftware/ccs/CCSCategoryNames_FullLabels.pdf

**Supplementary Table S2.** RxNorms and OMOP IDs used to identify opioid and gabapentin prescriptions.

| **Drug ingredient** | **RxNorm** | **OMOP Concept ID** |
|---|---|---|
| Hydrocodone | 5489 | 1174888 |
| Fentanyl | 4337 | 1154029 |
| Codeine | 2670 | 1201620 |
| Hydromorphone | 3423 | 1126658 |
| Meperidine | 6754 | 1102527 |
| Methadone | 6813 | 1103640 |
| Morphine | 7052 | 1110410 |
| Oxycodone | 7804 | 1124957 |
| Tramadol | 10689 | 1103314 |
| Gabapentin | 25480 | 797399 |

**Supplementary Table S3.** SNOMED codes used for opioid-related adverse outcome (OAO).

| Condition Name | SNOMED Code | OMOP Concept ID |
|---|---|---|
| Poisoning caused by opioid receptor agonist | 1148649003 | 606805 |
| Poisoning by opium alkaloid | 74264003 | 439223 |
| Poisoning by analgesic drug | 241748001 | 437158 |
| Opioid-induced sleep disorder | 88926005 | 4230779 |
| Opioid-induced organic mental disorder | 14784000 | 4032799 |
| Opioid dependence | 75544000 | 438120 |
| Harmful pattern of use of opioid | 5602001 | 438130 |
| Nondependent harmful pattern of use of opioid | 191909007 | 4099935 |

SNOMED concept identifiers were derived by mapping the ICD codes reported in Hajouji et al. (2023)* to standard OMOP concept IDs within the OMOP Common Data Model. All descendant concepts of these mapped codes were included in the definition of the opioid-related adverse outcome.

*El Hajouji O, Sun RS, Zammit A, Humphreys K, Asch SM, Carroll I, Curtin CM, Hernandez-Boussard T. Prediction of opioid-related outcomes in a medicaid surgical population: evidence to guide postoperative opiate therapy and monitoring. PLoS Computational Biology. 2023 Aug 14;19(8):e1011376.

**Supplementary Table S4.** Clinical code mappings and OMOP concept IDs used to identify comorbidity covariates.

| Clinical Condition | ICD-10 Code | OMOP Standard Concept ID |
|---|---|---|
| Essential hypertension, unspecified | I10.9 | 320128 |
| Neuropathy | G60.9 | 4301699 |
| Nephropathy | N14.2 | 4126119 |
| Retinopathy | H35.00 | 378416 |
| Lipid storage disorder, unspecified* | E75.6 | 4027782 |
| Hyperlipidemia, unspecified* | E78.5 | 432867 |
| Pure hypercholesterolemia, unspecified* | E78.00 | 437827 |
| Lipoprotein deficiency* | E78.6 | 435516 |
| Pure hyperglyceridemia* | E78.1 | 440360 |
| Chronic obstructive pulmonary disease, unspecified (COPD) | J44.9 | 255573 |
| Disorder of thyroid, unspecified | E07.9 | 141253 |
| Liver disease, unspecified | K76.9 | 194984 |
| Nerve disorder | E13.40 | 443730 |
| Cancer | — | 443392 |
| Depression disorder | — | 440383 |
| Anxiety disorder | — | 441542 |
| Alcohol disorder | — | 36714559, 3654404, 4214950 |
| Tobacco use | — | 903654, 903652, 4036084, 4005823, 4144271, 4038731, 4038735 |

* All these conditions were classified as a single covariate called lipid disorder.

Covariates were defined using standard OMOP condition concept identifiers. All descendant concepts in the OMOP concept hierarchy were included. Conditions were identified from the condition_occurrence table prior to the admission date.

**Supplementary Table S5.** Clinical code mappings and OMOP concept IDs used to identify comorbidity covariates

| **Laboratory Measurement** | **LOINC Group Code** | **OMOP Standard Concept ID** |
|---|---|---|
| Alanine aminotransferase (ALT), U/L | LG5272-2 | 40652525 |
| | LP382703-9 | 37047736 |
| Aspartate aminotransferase (AST), U/L | LG6033-7 | 40652640 |
| | LP382836-7 | 37059000 |
| Hemoglobin A1c (A1C) | — | 4184637 |
| | — | 3004410 |
| | — | 3005131 |
| | — | 3005673 |
| | — | 40762352 |
| | — | 3034639 |
| | — | 4197971 |
| | — | 3007263 |
| | — | 3003309 |

Laboratory covariates were defined using standard OMOP measurement concepts. The most recent value prior to the admission date was extracted from the measurement table. For A1C, multiple measurement concepts with varying reported units were included; all values were harmonized and converted to DCCT-aligned percentage (%) units (Diabetes Control and Complications Trial standard). Details of unit harmonization procedures are available in the study GitHub repository.

**Supplementary Table S6.** Performances of models using thresholds achieving the best F1 score.

| Model | Metric | Outcome | | | | Average |
|---|---|---|---|---|---|---|
| | | POU | Readmission | CP | OAO | |
| HCF-Transformer | Best F1 Threshold | 0.304 | 0.142 | 0.210 | 0.135 | |
| | Accuracy | 0.700 | 0.787 | 0.834 | 0.965 | 0.822 |
| | Precision | 0.649 | 0.202 | 0.323 | 0.264 | 0.360 |
| | Recall | 0.810 | 0.441 | 0.555 | 0.556 | 0.590 |
| | F1-score | 0.721 | 0.277 | 0.408 | 0.358 | 0.441 |
| FT-Transformer | Best F1 Threshold | 0.355 | 0.125 | 0.184 | 0.144 | |
| | Accuracy | 0.703 | 0.746 | 0.816 | 0.959 | 0.806 |
| | Precision | 0.658 | 0.179 | 0.301 | 0.245 | 0.346 |
| | Recall | 0.789 | 0.487 | 0.594 | 0.654 | 0.631 |
| | F1-score | 0.717 | 0.262 | 0.400 | 0.356 | 0.434 |
| ResNet | Best F1 Threshold | 0.396 | 0.092 | 0.275 | 0.18 | |
| | Accuracy | 0.724 | 0.732 | 0.825 | 0.960 | 0.810 |
| | Precision | 0.695 | 0.176 | 0.311 | 0.242 | 0.356 |
| | Recall | 0.751 | 0.514 | 0.568 | 0.596 | 0.607 |
| | F1-score | 0.722 | 0.262 | 0.402 | 0.344 | 0.432 |
| Random Forest | Best F1 Threshold | 0.358 | 0.117 | 0.178 | 0.129 | |
| | Accuracy | 0.707 | 0.727 | 0.817 | 0.967 | 0.805 |
| | Precision | 0.662 | 0.176 | 0.306 | 0.271 | 0.354 |
| | Recall | 0.788 | 0.527 | 0.607 | 0.529 | 0.613 |
| | F1-score | 0.720 | 0.263 | 0.407 | 0.358 | 0.437 |

**Supplementary Table S7.** Average differences in total relative risk (TRR) between model-recommended and actual opioids prescribed at discharge, including mean differences and 95% confidence intervals (CI), highlighting the preferences determined by the HCF-Transformer model.

| Model-recommended Opioid | Actual Opioid | Count | % | TRR Mean Difference | SD | ± 95% CI |
|---|---|---|---|---|---|---|
| Tramadol | Hydrocodone | 6723 | 21.3 | 0.07825 | 0.02615 | 0.00063 |
| Tramadol | Oxycodone | 5948 | 18.8 | 0.0429 | 0.00973 | 0.00025 |
| Tramadol | Non-opioid | 5690 | 18.0 | 0.39903 | 0.12738 | 0.00331 |
| Hydrocodone | Non-opioid | 3747 | 11.9 | 0.47628 | 0.22625 | 0.00724 |
| Hydrocodone | Oxycodone | 2845 | 9.0 | 0.12489 | 0.08625 | 0.00317 |
| Hydrocodone | Hydrocodone | 1904 | 6.0 | 0 | 0 | 0 |
| Codeine | Hydrocodone | 1242 | 3.9 | 0.05929 | 0.04053 | 0.00225 |
| Codeine | Oxycodone | 1129 | 3.6 | 0.09208 | 0.04906 | 0.00286 |
| Tramadol | Codeine | 528 | 1.7 | 0.09926 | 0.06597 | 0.00563 |
| Tramadol | Tramadol | 514 | 1.6 | 0 | 0 | 0 |
| Codeine | Non-opioid | 454 | 1.4 | 0.30159 | 0.06813 | 0.00627 |
| Hydrocodone | Tramadol | 189 | 0.6 | 0.18031 | 0.25412 | 0.03623 |
| Tramadol | Hydromorphone | 117 | 0.4 | 0.31678 | 0.0808 | 0.01464 |
| Hydrocodone | Hydromorphone | 98 | 0.3 | 0.55118 | 0.22502 | 0.04455 |
| Hydrocodone | Codeine | 96 | 0.3 | 0.13918 | 0.13077 | 0.02616 |
| Hydrocodone | Morphine | 78 | 0.2 | 0.71231 | 0.34273 | 0.07606 |
| Codeine | Codeine | 73 | 0.2 | 0 | 0 | 0 |
| Codeine | Tramadol | 73 | 0.2 | 0.07064 | 0.0581 | 0.01333 |
| Tramadol | Morphine | 44 | 0.1 | 0.40089 | 0.11516 | 0.03403 |
| Oxycodone | Oxycodone | 23 | 0.1 | 0 | 0 | 0 |
| Codeine | Hydromorphone | 20 | 0.1 | 0.39443 | 0.12699 | 0.05565 |
| Oxycodone | Hydrocodone | 19 | 0.1 | 0.01951 | 0.01158 | 0.00521 |
| Oxycodone | Non-opioid | 8 | 0.0 | 0.445 | 0.14164 | 0.09815 |
| Oxycodone | Codeine | 4 | 0.0 | 0.06248 | 0.02227 | 0.02183 |
| Codeine | Morphine | 3 | 0.0 | 0.34077 | 0.1449 | 0.16397 |
| Oxycodone | Hydromorphone | 1 | 0.0 | 0.229 | NA | NA |
| Oxycodone | Tramadol | 1 | 0.0 | 0.0166 | NA | NA |

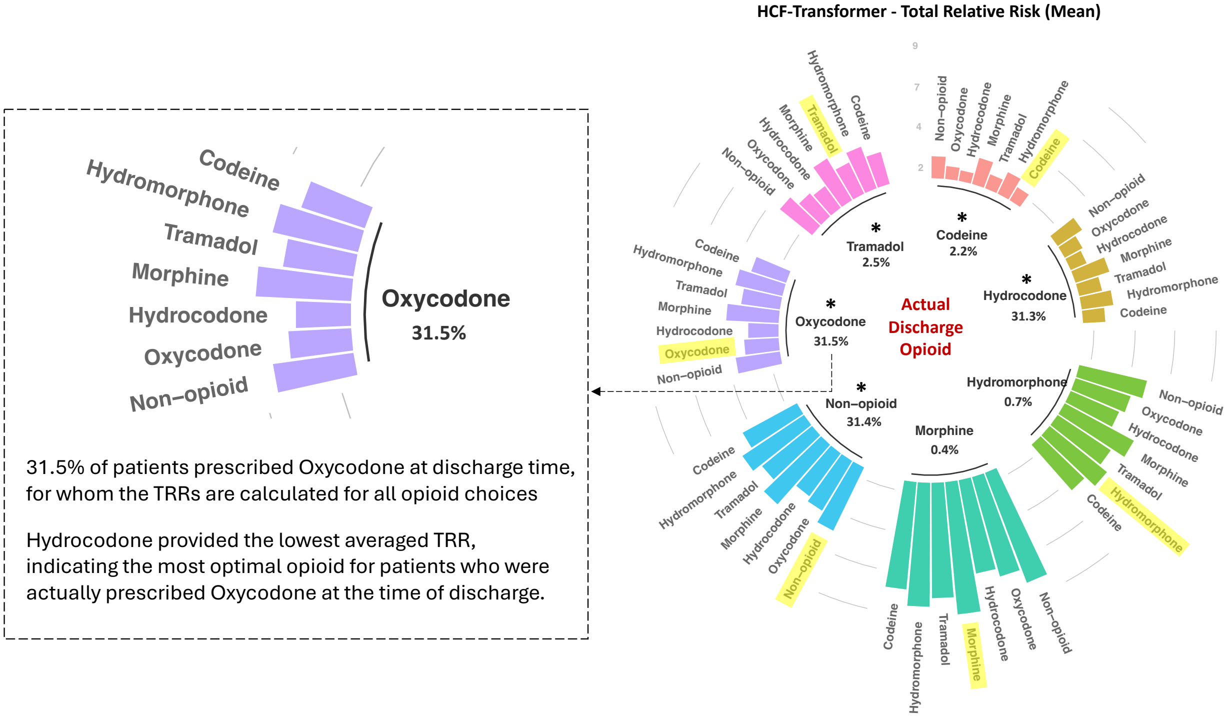


**Supplementary Figure S1.** Total relative risks (TRR) of actual vs. alternative discharge opioids based on the HCF-Transformer model obtained from thresholds that align closely with the outcome prevalences shown in Table 1.

* indicates ANOVA $p < .01$ with effect size eta-squared > .01. Highlighted opioids represent TRRs for the actual prescribed discharge opioid.
